# Portable, Low-Field Magnetic Resonance Imaging Sensitively Detects and Accurately Quantifies Multiple Sclerosis Lesions

**DOI:** 10.1101/2022.03.11.22272272

**Authors:** T. Campbell Arnold, Danni Tu, Serhat V. Okar, Govind Nair, Samantha By, Karan Kawatra, Timothy E. Robert-Fitzgerald, Lisa M. Desiderio, Matthew K. Schindler, Russell T. Shinohara, Daniel S. Reich, Joel M. Stein

## Abstract

Magnetic resonance imaging is a fundamental tool in the diagnosis and management of neurological diseases such as multiple sclerosis (MS). New portable, low-field MRI scanners could potentially lower financial and technical barriers to neuroimaging and reach underserved or disabled populations. However, the sensitivity of low-field MRI for MS lesions is unknown. We sought to determine if white matter lesions can be detected on a 64mT low-field MRI, compare automated lesion segmentations and total lesion burden between paired 3T and 64mT scans, and identify features that contribute to lesion detection accuracy. In this prospective, cross-sectional study, same-day brain MRI (FLAIR, T1, and T2) scans were collected from 36 adults (32 women; mean age, 50 ± 14 years) with known or suspected MS using 3T (Siemens) and 64mT (Hyperfine) scanners at two centers. Images were reviewed by neuroradiologists. MS lesions were measured manually and segmented using an automated algorithm. Statistical analyses assessed accuracy and variability of segmentations across scanners and systematic scanner biases in automated volumetric measurements. Lesions were identified on 64mT scans in 94% (31/33) of patients with confirmed MS. The smallest lesions manually detected were 5.7 ± 1.3 mm in maximum diameter at 64mT vs 2.1 ± 0.6 mm at 3T. Automated lesion burden estimates were highly correlated between 3T and 64mT scans (r = 0.89, p < 0.001). Bland-Altman analysis identified bias in 64mT segmentations (mean = 1.6 ml, standard error = 5.2 ml, limits of agreement = -19.0–15.9 ml), which over-estimated low lesion burden and under-estimated high burden (r = 0.74, p < 0.001). Visual inspection revealed over-segmentation was driven by flow-related hyperintensities in veins on 64mT FLAIR. Lesion size drove segmentation accuracy, with 93% of lesions >1.0 ml and all lesions >1.5 ml being detected. These results demonstrate that in established MS, a portable 64mT MRI scanner can identify white matter lesions, and disease burden estimates are consistent with 3T scans.

**Highlights:**

- Paired, same-day 3T and 64mT MRI studies were collected in 36 patients
- 64mT MRI showed 94% sensitivity for detecting any lesions in established MS cases
- The diameter of the smallest detected lesion was larger at 64mT compared to 3T
- Disease burden estimates were strongly correlated between 3T and 64mT scans
- Low-field MRI can detect white matter lesions, though smaller lesions may be missed

## 1. Introduction

Multiple sclerosis (MS) is a complex inflammatory and degenerative disease of the central nervous system (1). MS causes demyelinating lesions, typically assessed using magnetic resonance imaging (MRI). Imaging features related to white matter lesions (WML), such as number, volume, and dissemination in space and time, are key diagnostic criteria of MS (2) and determine treatment courses and clinical trial eligibility (3). Early diagnosis leads to better clinical outcomes, including delayed disease progression and reduced severity (4).

Although MS affects ∼800,000 people in the United States (5) and likely >2.5 million people globally (6), the significant cost, infrastructure, and technical requirements associated with traditional high-field-strength MRI limit access to imaging worldwide (7). The scarcity is particularly felt in low-resource, sparsely populated, and rural areas (8). As the lack of diagnostic imaging can lead to delayed diagnosis and treatment, which result in worsening health disparities (9), there is renewed interest in low-field MRI, which employs magnets with a field strength of 1 tesla (T) or lower, as a lower-cost and potentially portable alternative to high-field MRI (10).

Recent improvements in hardware as well as image reconstruction and processing algorithms (11) have made low-field MRI promising in contexts where modest resolution is sufficient for diagnostic purposes (12). The clinical utility of portable low-field MRI has already been demonstrated for bedside monitoring in intensive care settings, where patients may not be stable enough to transport for traditional imaging (13–15). In the outpatient treatment of diseases such as MS, portable low-field MRI has the potential to lower barriers to accessing MRI technology and allow more frequent monitoring of disease activity (16), however its sensitivity and accuracy have not been explored. In particular, WML to background signal intensities and size thresholds for detection are unknown.

In this study, we assessed the feasibility of portable low-field MRI for MS lesion identification and lesion burden estimation. We collected paired same-day brain MRI from adults with MS at 3T and 64mT at two different institutions and compared lesion detection using both manual and automated measurements. We anticipated that MS lesions would be detectable at 64mT, though sensitivity to small lesions would likely be reduced. Finally, we explored a simple approach for super-resolution imaging of small lesions based on multi-acquisition image averaging.

## 2. Materials and Methods

### 2.1 Participants & Imaging

Among adult outpatients undergoing clinical 3T brain MRI for known or suspected MS between October 2020 and April 2021, 36 patients (Fig. 1) were recruited at site A (N=21) and site B (N=15). All patients received same-day 3T and 64mT MRI. Demographic information was collected from clinical notes and included age, sex, race, clinical phenotype, disease duration, Expanded Disability Status Scale (EDSS), and current disease modifying therapy (Table 1). This study was approved by each site’s institutional review board, and patients provided written, informed consent.

**Table 1.** Patient Demographics. Demographic information and clinical history for 36 consecutive MS patients included in the study. An asterisk indicates a significant difference between sites. Abbreviations: Expanded disability status scale (EDSS), relapsing-remitting multiple sclerosis (RRMS), primary progressive multiple sclerosis (PPMS), and secondary progressive multiple sclerosis (SPMS), clinically isolated syndrome (CIS), radiologically isolated syndrome (RIS), neuromyelitis optica (NMO), idiopathic transverse myelitis (ITM).

|  | Total (N = 36) | Site A (N = 21) | Site B (N = 15) |
| --- | --- | --- | --- |
| Age (years) | 49.6 ± 14.2 | 45.3 ± 13.6 * | 55.7 ± 12.7 * |
| Sex (women/men) | 32/4 | 19/2 | 13/2 |
| Race/ethnicity (White/Black/Hispanic) | 26/9/1 | 14/7/0 | 12/2/1 |
| Disease duration (years) | 13.7 ± 11.2 | 10.2 ± 9.6 * | 18.5 ± 11.5 * |
| EDSS (0–10) | 1.5 (IQR = 2.0) | 1.5 (IQR = 2.0) | 2.0 (IQR = 1.25) |
| Phenotype |  | RRMS (18), CIS (1), NMO (1), RIS (1) | RRMS (10), SPMS (2), CIS (1), ITM (1), PPMS (1) |
| Current disease modifying therapy |  | ocrelizumab (9), natalizumab (2), other (6), none (4) | dimethyl fumarate (6), ocrelizumab (3), other (3), none (6) |

**Figure 1.**
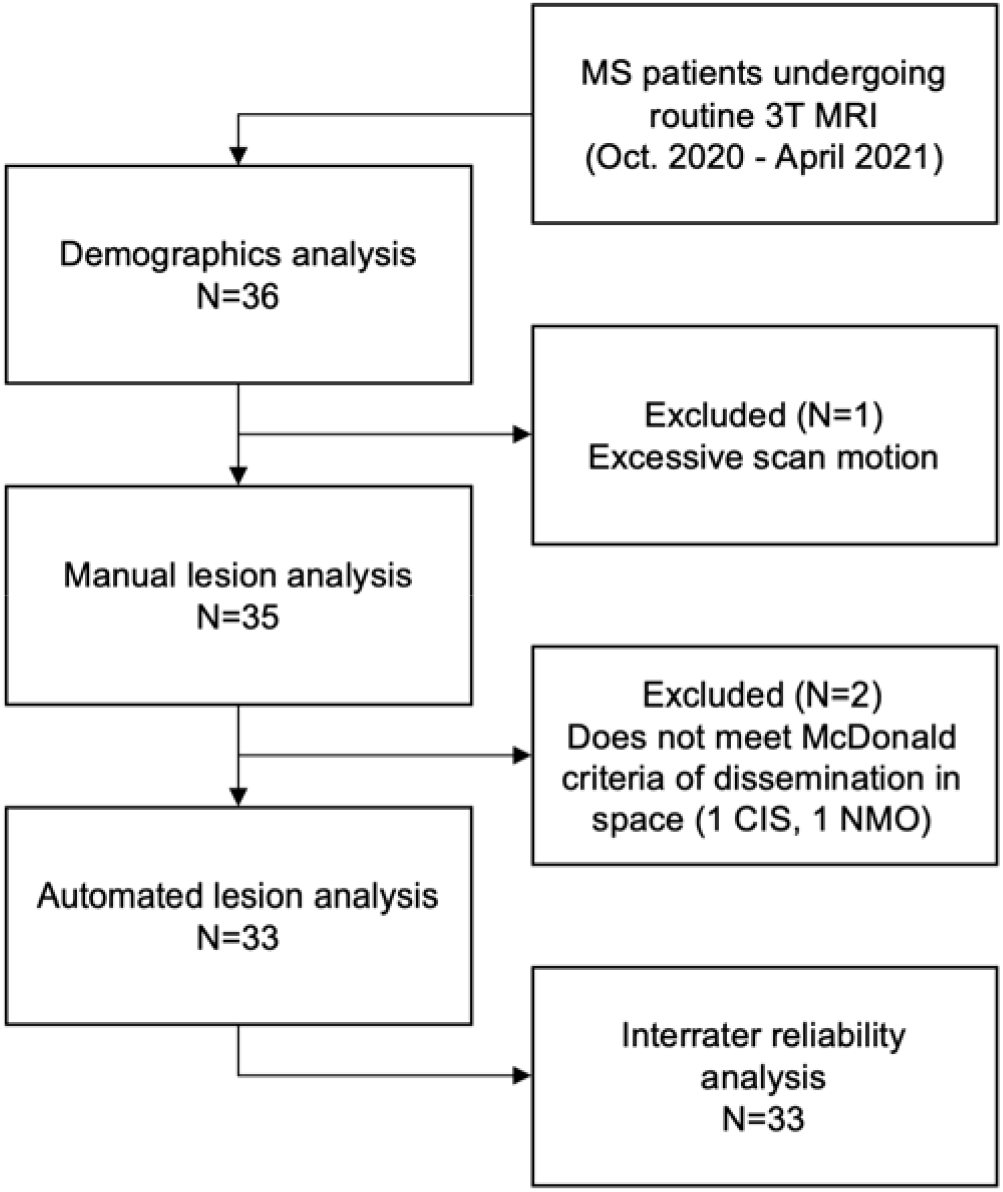
Flow chart of study participants. Abbreviations: multiple sclerosis (MS), clinically isolated syndrome (CIS), neuromyelitis optica (NMO).

High-field MRI was performed on 3T scanners (Siemens, Erlangen, Germany). Each site used a standardized, whole-brain imaging protocol, which included 3D T1-weighted (T1w), T2-weighted (T2w), and 3D T2-FLAIR sequences (Fig. 2A). Sequence parameters are listed in Table 2. At site B, patients received gadolinium (gadobutrol, 0.1 mmol/L) prior to 3T scanning; 64mT scans were obtained after contrast-enhanced 3T scans with mean post-gadolinium duration of 58 ± 21 minutes.

**Table 2.** Sequence parameters for study scans. Abbreviations: Tesla (T), T1-weighted (T1w), T2-weighted (T2w), Fluid-attenuated inversion recovery (FLAIR), echo time (TE), repetition time (TR), inversion time (TI).

| Sequence | Site | Field Strength (T) | TE (ms) | TR (s) | TI (s) | Resolution (mm) | Scan-time (min:sec) | Averages |
| --- | --- | --- | --- | --- | --- | --- | --- | --- |
| T1w-MPRAGE | A | 3 | 2.48 | 1.9 | 0.9 | 1.0x1.0x1.0 | 4:18 | 1 |
| T1w-MP2RAGE | B | 3 | 2.92 | 5 | 0.7, 2.5 | 1.0x1.0x1.0 | 8:30 | 1 |
| T1w | Both | 0.064 | 6.26 | 1.5 | 0.3 | 1.5x1.5x5 | 4:52 | 1 |
| T2-FLAIR 3D | A | 3 | 398 | 5 | 1.6 | 1.0x1.0x1.0 | 5:02 | 1 |
| T2-FLAIR 3D | B | 3 | 352 | 4.8 | 1.8 | 1.0x1.0x1.0 | 7:15 | 1 |
| T2-FLAIR | Both | 0.064 | 200 | 4 | 1.4 | 1.6x1.6x5 | 9:29 | 1 |
| T2w | A | 3 | 103 | 5.5 | N/A | 0.5x0.5x5.2 | 1:46 | 2 |
| T2w | B | 3 | 82 | 5 | N/A | 0.34x0.34x3.0 | 4:30 | 1 |
| T2w | Both | 0.064 | 209 | 2 | N/A | 1.5x1.5x5 | 7:01 | 1 |

**Figure 2.**
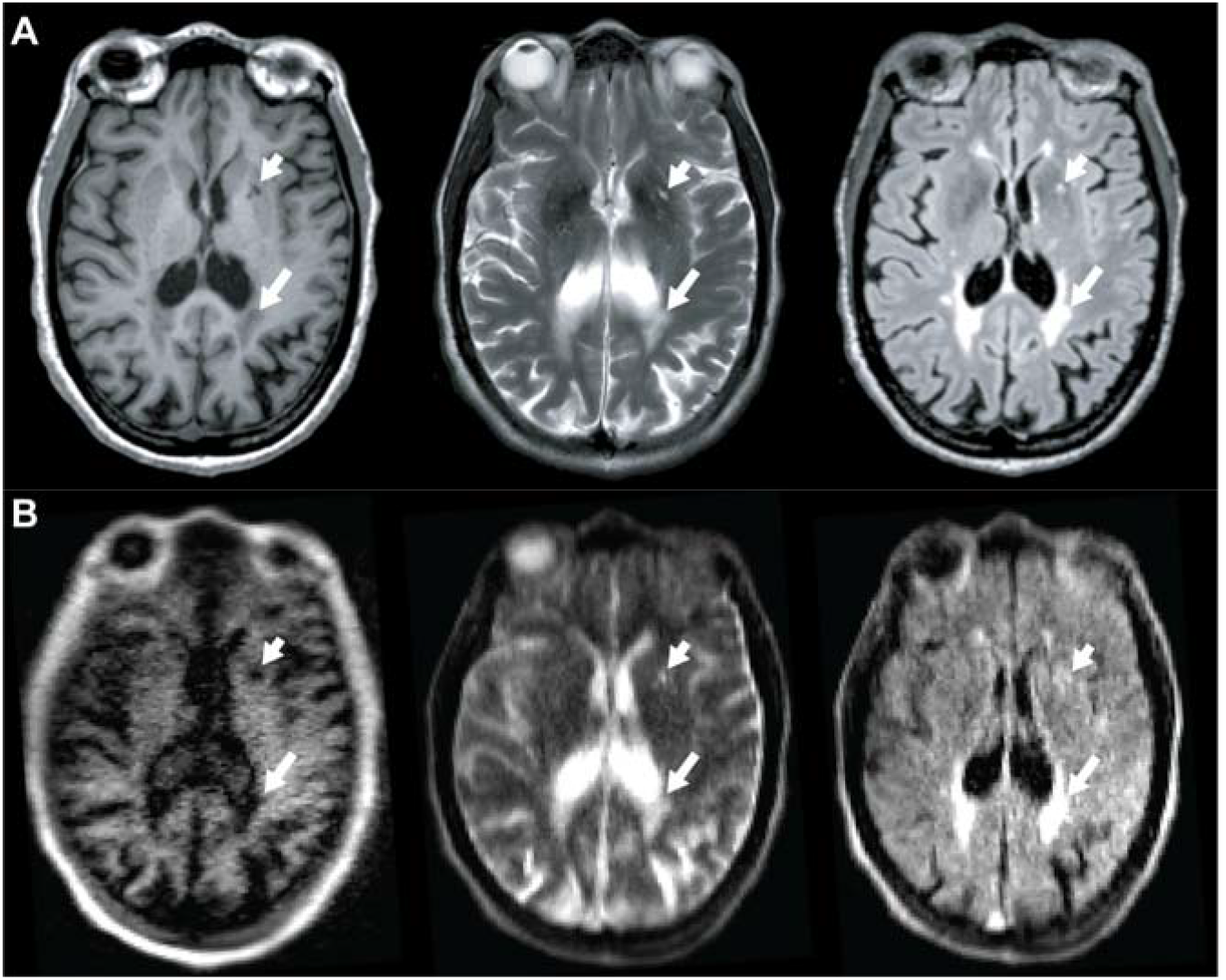
MS lesions appear similar on 3T and 64mT pulse sequences. Paired 3T (A) and 64mT (B) images from a 66-70 year-old female with stable RRMS showing deep gray matter lesions (short arrow) and periventricular white matter lesions (long arrow) on both scanners. Sequences include T1-weighted (left), T2-weighted (center), and T2-FLAIR (right).

Same-day, low-field MRI was performed on portable 64mT Swoop MRI systems (Hyperfine, Guilford, CT). Whole-brain, 3D T1w, T2w, and T2-FLAIR scans, analogous to those acquired at 3T, were collected (Fig. 2B). Including localizer and pre-scan calibration, total scan time was 26:06 minutes.

### 2.2 Manual Review and Lesion Measurement

MRI scans were reviewed for WML by two neuroradiologists (DSR and JMS, 19 and 8 years experience). Maximum diameters (Dmax) of the smallest and largest WML visually detectable at each field strength were manually measured by a neuroradiologist (JMS) and a neurologist (SVO) with MS MRI expertise (3 years experience) using ITK-SNAP (17). All measurements were made on T2-FLAIR scans. Lesions were assessed in axial planes as well as sagittal and coronal reformations, and Dmax measurements were made on the plane with the largest lesion diameter. In confluent periventricular lesions, Dmax was measured perpendicular to the ventricle. Low-field imaging was evaluated prior to 3T scans to avoid interpretation bias, and image sets were reviewed separately. Inter-rater reliability was assessed using two-way random, single-measure intraclass correlation coefficients (ICC) with 95% confidence intervals (CI) reported. Patients that did not meet the McDonald criteria for dissemination of lesions in space were excluded from subsequent analyses (2).

### 2.3 Automated Lesion Segmentation

The same segmentation pipeline was applied to 3T and 64mT images. Images were preprocessed using N4 bias correction (18), and each T2-FLAIR volume was rigidly registered to the corresponding T1w volume using Advanced Normalization Tools (ANTs) (19,20). A brain mask was obtained using Multi-Atlas Skull-Stripping (MASS) (21). Finally, to enable comparisons across patients, image intensities were normalized using White Stripe (22) within each sequence. Lesion segmentation was performed using the Method for Inter-Modal Segmentation Analysis (MIMoSA) (23,24), an automated pipeline that leverages shared information (coupling) between modalities to produce probability maps of WML (Fig. S1). All probability maps were thresholded at 0.2 to generate binary lesion masks, manually selected based on prior empirical evidence.

### 2.4 Automated Segmentation Evaluation

Estimation of total lesion burden was the primary performance measure compared between 3T and 64mT. Two lesion burden estimates were obtained for each patient by calculating lesion segmentation volumes for the respective scanners. The relationship between volume estimates was assessed using Pearson’s correlation. Bland-Altman plots were used to determine agreement and assess for systematic scanner biases.

Similarity between segmentation masks was assessed using the Dice-Sørensen coefficient (Dice), which measures the overlap between two images (*X* and *Y*):

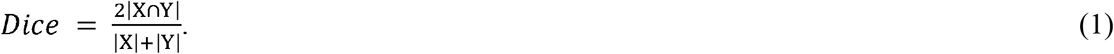

Dice scores range from 0–1, with 1 indicating perfect segmentation overlap. Prior to Dice calculation, 3T and 64mT images were coregistered using ANTs (20). While Dice score may not reflect segmentation quality when the number of target objects is not known *a priori*, this measure has been included as it is widely used and allows for comparisons across studies (25). All 3T and 64mT segmentations were manually reviewed to verify overlapping regions were WMLs rather than false positive detections.

### 2.5 Size and Intensity Analysis

Connected-components analysis was used to identify individual lesions in automated 3T and 64mT segmentations (26). Sensitivity to individual lesions at low-field MRI was assessed using the true-positive rate (TPR), or the proportion of lesions correctly identified:

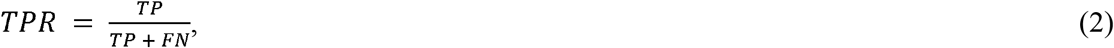

where true positives (*TP*) are defined as lesions where 64mT and 3T segmentations overlap and false negatives (*FN*) are defined as lesions with 3T segmentation but no 64mT segmentation overlap. The false-discovery rate (FDR) was assessed as:

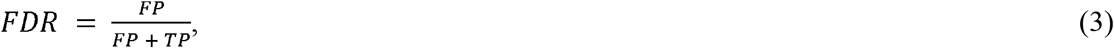

where false positives (*FP*) are defined as lesions with 64mT segmentation but no 3T segmentation overlap. Lesion overlap was defined as at least one shared voxel between the 3T and 64mT lesion segmentations. To understand the impact of lesion features on detection rates, TPR and FDR were plotted as a function of lesion size and normalized lesion intensity (22).

### 2.6 Super Resolution Imaging

Low-field MRI necessitates a trade-off between signal-to-noise ratio and image resolution, which limits the minimum detectable lesion size. However, image quality can be increased by taking advantage of partial volume effects in multiple scans (27). In one patient, multi-acquisition volume averaging was explored. Eight 64mT T2-FLAIR acquisitions (TE = 1.8 s, TR = 4 s, TI = 1.4 s, averages = 80, scan time = 6:03 min, resolution = 1.8×1.8×5 mm) were collected with head repositioning between scans (total scan time: 48:24 minutes). Images were resliced to 1 mm isotropic resolution with linear interpolation, underwent affine registration to the initial acquisition, and were averaged to create a single high-resolution image. Super resolution images were iteratively generated for each additional acquisition, for a total of eight images.

To quantify lesion conspicuity, we manually segmented the white matter lesion and a similarly sized region in normal appearing ipsilateral white matter on 3T imaging using ITK-SNAP (17). The 3T image and lesion segmentations were registered to the initial 64mT acquisition. We calculated lesion conspicuity as the ratio of the difference and the sum of mean intensity in the two segmentations:

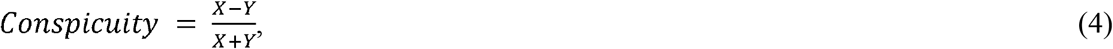

where *X* is the mean intensity in the lesion segmentation and *Y* is the mean intensity of the normal appearing white matter segmentation. The segmentations were applied to each super resolution iteration to calculate conspicuity for that iteration.

### 2.7 Statistics and Data/Code Availability

All code related to this study is publicly available. The MIMoSA algorithm is available in R as a Neuroconductor package (https://neuroconductor.org/package/details/mimosa) and on GitHub (https://github.com/avalcarcel9/mimosa/). T-tests, Pearson’s correlation, and summary statistics were calculated using scipy (v1.5.2) and numpy (v1.19.2) in Python (v3.8.5). Bland-Altman plots were visualized using pyCompare (v1.5.1) while boxplots and correlations utilized seaborn (v0.11.0). Inter-rater reliability was calculated using irr (v0.84.1) in R (v4.0.3). A manuscript companion containing all analyses is available on GitHub (https://github.com/penn-cnt/Arnold_LF-MRI_MS). The data generated in this study can be made available, with protected health information removed, upon reasonable request to the corresponding author and with a data sharing agreement between institutions in place.

## 3. Results

### 3.1 Patient Demographics

We collected data from 36 adults with known or suspected MS. The patient population had a mean age of 49.6 (SD: 14.2) years and was composed of 32 women and 4 men (Table 1). The mean duration of disease was 13.7 years (SD: 11.2), and patients had a median EDSS of 1.5 (interquartile range = 2). Patients from site B were significantly older than those from site A (two-sample t-test, t=2.2, p = 0.03, site A: 45.3 years old, site B: 55.7 years old) and had a correspondingly longer duration of disease (two-sample t-test, t=2.3, p = 0.03, site A: 10.2 years, site B: 18.5 years). Additional demographic information is provided in Table 1. After initial scan review, three patients were excluded from further analysis: One patient had excessive motion in the scanner and two patients did not meet the MS diagnostic criteria of having lesion dissemination in space (DIS) (2). All subsequent analyses are based on the remaining 33 patients (Fig. 1).

### 3.2 Manual Measurements

MS lesions on 64mT are characterized by T1w hypointensity and T2w/T2-FLAIR hyperintensity, similar to 3T imaging (Fig. 2). At 64mT, lesions were identified by at least one rater in 94% (31/33) of patients with confirmed lesions on 3T imaging. In one patient, only one rater identified lesions at 64mT; all other low-field ratings were concordant. The largest and smallest lesions in each image were identified, and the Dmax was recorded. The 64mT scanner showed 100% sensitivity for detecting WML when there was at least one lesion with Dmax >5 mm (31/33 patients, 94%). Across patients, there was no significant difference in Dmax for the largest lesions measured at 64mT (15.1 ± 5.9 mm) and 3T (14.8 ± 6.6 mm) (Fig. 3A). However, the mean Dmax for the smallest WML detectable was significantly larger (paired t-test, t=19.6, p < 0.001) on 64mT (5.7 ± 1.3 mm) compared to 3T (2.1 ± 0.6 mm) (Fig. 3B). There was no effect of scan site on Dmax measurements, however there was a significant difference between rater 1 (2.3 ± 0.5 mm) and rater 2 (1.9 ± 0.6 mm) for the smallest lesion detected at 3T (paired t-test, t=4.8, p < 0.001). No gadolinium enhancing lesions were seen on 3T or 64mT imaging.

**Figure 3.**
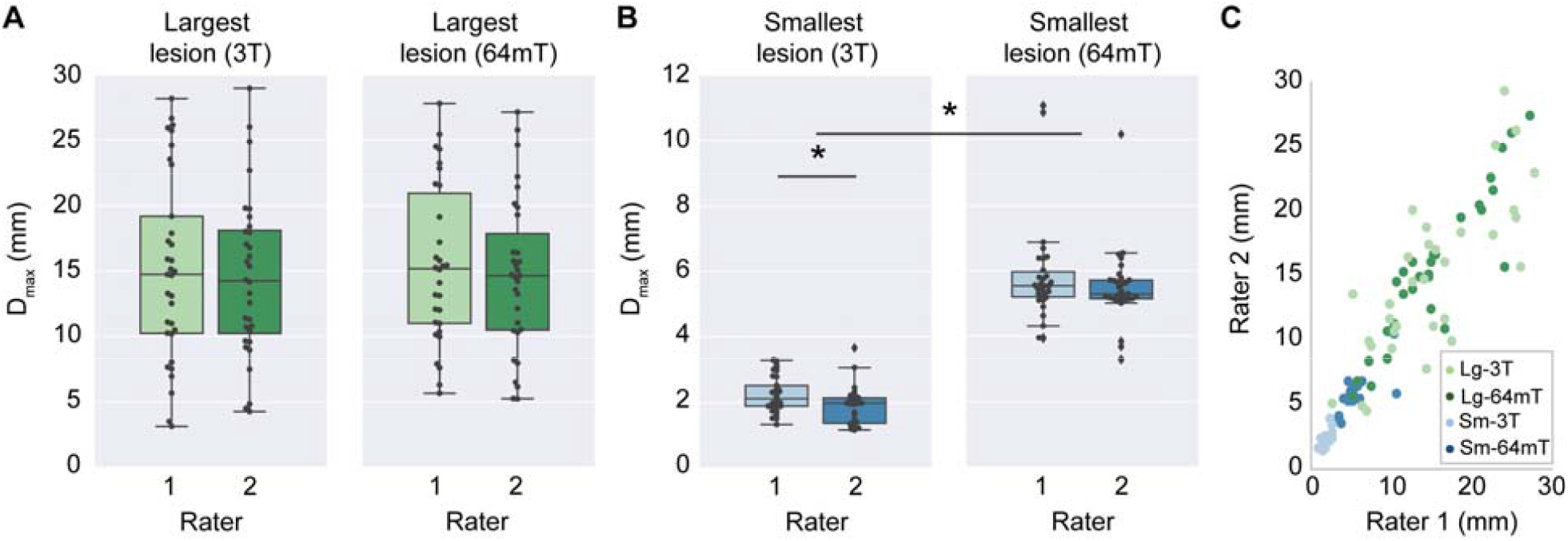
Manual lesion size measurements and interrater reliability. Raters from each site independently measured the maximum diameter (Dmax) of the smallest lesion (Sm) and largest lesion (Lg) in 3T and 64mT imaging for all patients. (A) For the largest lesion measurements, there were no significant differences between raters at 3T (t = 1.3, p = 0.19) or 64mT (t = 1.2, p = 0.23); additionally, there was no difference between 3T and 64mT measurements (t = 0.04, p = 0.97). (B) For the smallest lesion measurements, there was a significant difference between raters for 3T measurements (t = 4.83, p < 0.001) although 64mT measurements were not significantly different (t = 1.67, p = 0.11); additionally, the diameter of the smallest lesion was significantly lower (t = 19.6, p < 0.001) when measured on 3T (mean 2.1 mm) compared to 64mT (mean 5.7 mm). (C) Across all lesions there was a strong correlation (r = 0.90, p < 0.001) between raters. There was significant intraclass correlation for the largest lesion at 3T (ICC = 0.77, CI = [0.58-0.88]), largest lesion at 64mT (ICC = 0.91, CI = [0.83-0.96]), smallest lesion at 3T (ICC = 0.62, CI = [0.12-0.83]), and smallest lesion at 64mT (ICC = 0.66, CI = [0.4-0.82]), indicating a high degree of agreement between rater measurements for both 3T and 64mT.

### 3.3 Interrater Reliability

The smallest and largest lesion in each image were independently measured by two raters to assess interrater reliability (Fig. 3). The ICC for each patient’s largest lesion measured at 3T and 64mT was 0.77 (CI = [0.58-0.88]) and 0.91 (CI = [0.83-0.96]) respectively, indicating high interrater reliability for large lesions on both scanners (Fig. 3A). Similarly, when measuring each patient’s smallest lesion there was a significant relationship between raters at both 3T (ICC = 0.62, CI = [0.12-0.83]) and 64mT (ICC = 0.66, CI = [0.4-0.82]) (Fig. 3B). This indicates that measurements made by raters had a similar degree of reliability at 3T and 64mT. Of note, the average smallest lesions detected (3T: 2.1 ± 0.6 mm, 64mT: 5.7 ± 1.3 mm) approached the respective resolution limits for each system (3T: 1 mm, 64mT: 5 mm).

### 3.4 Total Lesion Burden Estimates

To obtain more objective measures, 3T and 64mT image sets were processed with the same automated lesion segmentation algorithm. Initial qualitative review of segmentation overlays revealed similar patterns of lesion segmentation, particularly with respect to large periventricular lesions (Fig. 4). Quantitative comparisons indicated that estimates of total lesion burden were highly correlated (r = 0.89, p < 0.001) (Fig. 5A). Mean lesion burden estimates were not significantly different (paired-t-test, t = 1.0, p = 0.32) between 3T (11.9 ± 16.5 ml) and 64mT (13.5 ± 10.2 ml) images.

**Figure 4.**
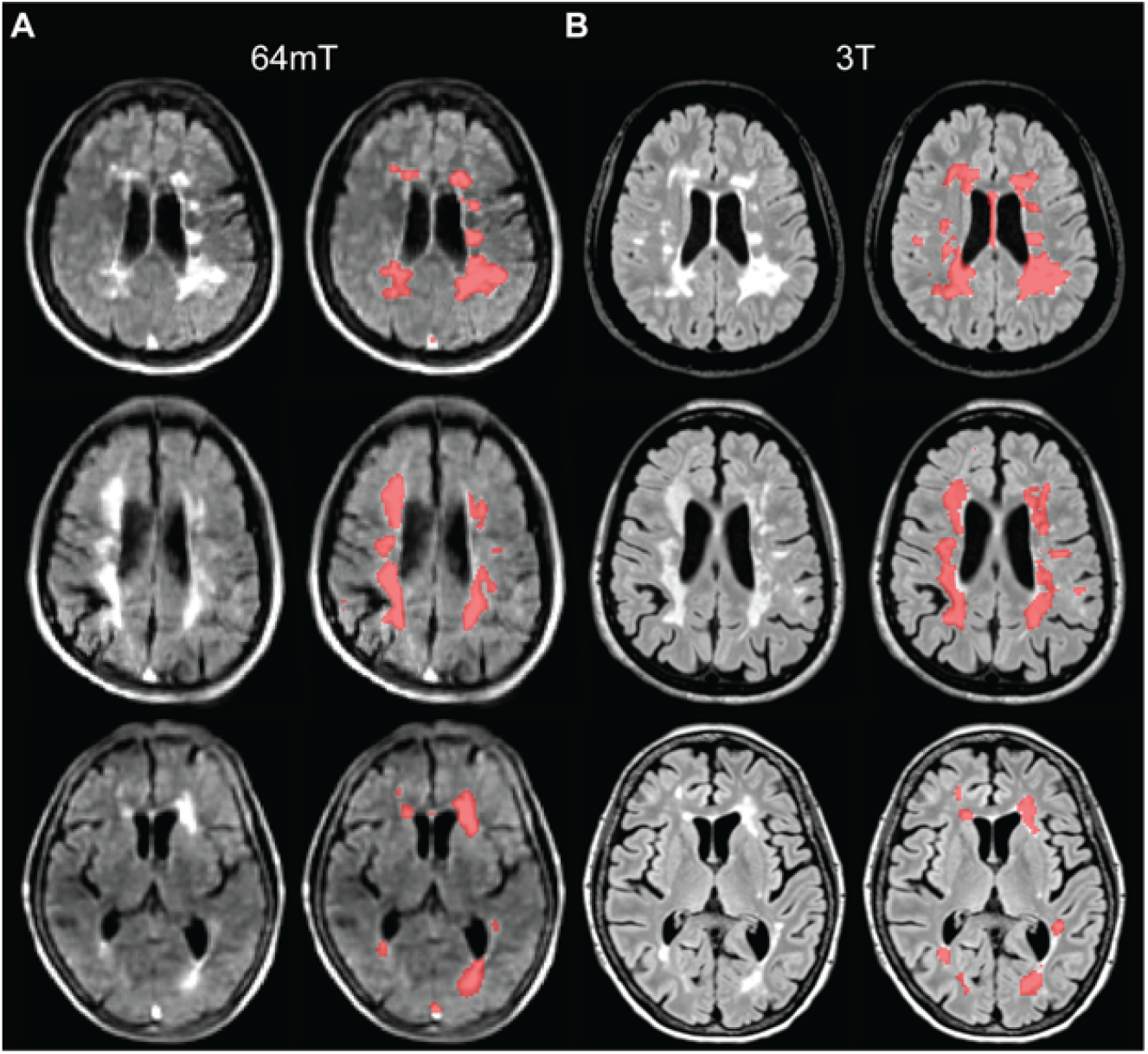
Automated lesion segmentations at 3T and 64mT overlap. (A) 64mT FLAIR images for three cases (left) with automated lesion segmentations generated from the 64mT images using MIMoSA overlaid (right). (B) Corresponding 3T FLAIR images for the same three cases (left) with 3T based segmentations (right). Patients from top to bottom are a 51-55 year-old female with RRMS, 41-45 year-old female with RRMS, and 71-75 year-old female with RRMS. All images were coregistered to 64mT T1-weighted images for comparison. Segmentations generated from 64mT and 3T scanners show similar patterns.

**Figure 5.**
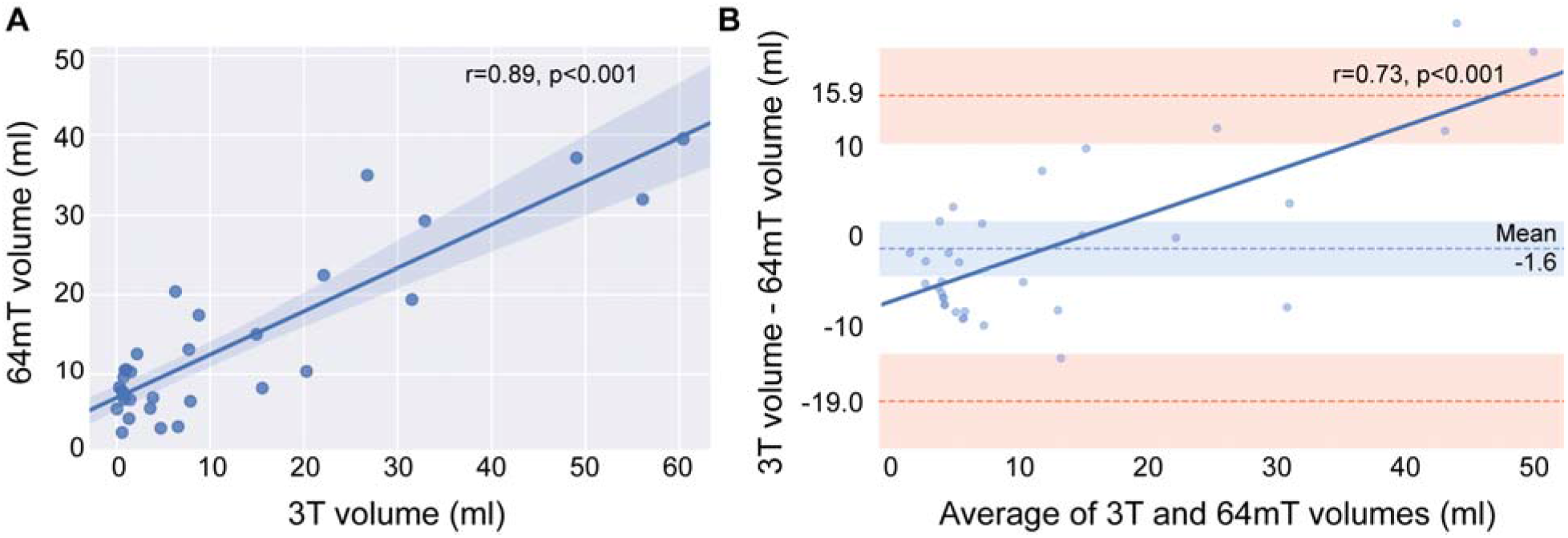
Total lesion volume measured at 3T and 64mT shows agreement. (A) Pearson’s correlation (r = 0.89, p < 0.001) and (B) Bland-Altman plot showing agreement between 3T and 64mT segmentation volumes (bias -1.6 ml, standard error of measurement = 5.2 ml, 95% limit of agreement -19.0 to 15.9 ml). Pearson’s correlation (r = 0.74, p < 0.001) in dark blue indicates over-segmentation at 64mT when lesion burden is low and under-segmentation when burden is high.

A Bland-Altman plot for agreement between 3T and 64mT lesion burden estimates is presented in Figure 5B. The mean difference was 1.6 ml with a 5.2 ml standard error of measurement, and the 95% limits of agreement were -19.0 to 15.9 ml. There was a significant correlation (r = 0.74, p < 0.001) between pairwise differences and averages, indicating that compared to 3T, the 64mT segmentations overestimate low lesion burdens and underestimate high lesion burdens. Visual inspection revealed that false positives contributing to over-segmentation were predominantly due to flow-related high signal intensity in veins, hyperintensity in non-lesion structures (such as the pineal gland), and areas of artifactual peripheral high signal in cortical/subcortical tissue on 64mT FLAIR sequences (Fig. S2).

### 3.5 Automated Segmentation Overlap

Across patients, there was a wide range in overlap between 3T and 64mT segmentation pairs (Dice: mean = 0.23, standard deviation = 0.21, max = 0.65, min = 0), with automated segmentations overlapping in 91% (30/33) of patients. Two patients had no segmentation overlap and one patient was excluded because the overlapping region was a hyperintense pineal gland, not a WML. All three patients without lesion overlap were in the bottom 12% of total lesion burden. To characterize the full range of segmentation quality across the dataset, Figure S3 illustrates segmentations from each quartile of the Dice distribution. Larger lesion size is frequently associated with higher Dice (25). We found in our dataset that total lesion burden at 3T was highly correlated with Dice (r = 0.81, p < 0.001) (Fig. S4).

### 3.6 Lesion Sensitivity and False Discovery

In each segmentation, individual lesions were identified using connected-components analysis (26). For each lesion, volume and mean intensity were quantified. The true-positive rate (TPR) and false-discovery rate (FDR) were calculated across a range of lesion size and intensity thresholds (Fig. 6). The TPR increases dramatically with lesion size, reaching 93% for lesions >1 ml and 100% for lesions >1.5 ml. The FDR decreases with lesion size, reaching 36% for lesions >1 ml, 22% for lesions >1.5ml, and 3% for lesions >2.5 ml. TPR also increases with mean lesion intensity, indicating that lesion intensity influences sensitivity; however, FDR remains high (>75%) regardless of the sensitivity, indicating a large number of false positive detections across intensity thresholds. Examples of false positive detections can be seen in Figure S2.

**Figure 6.**
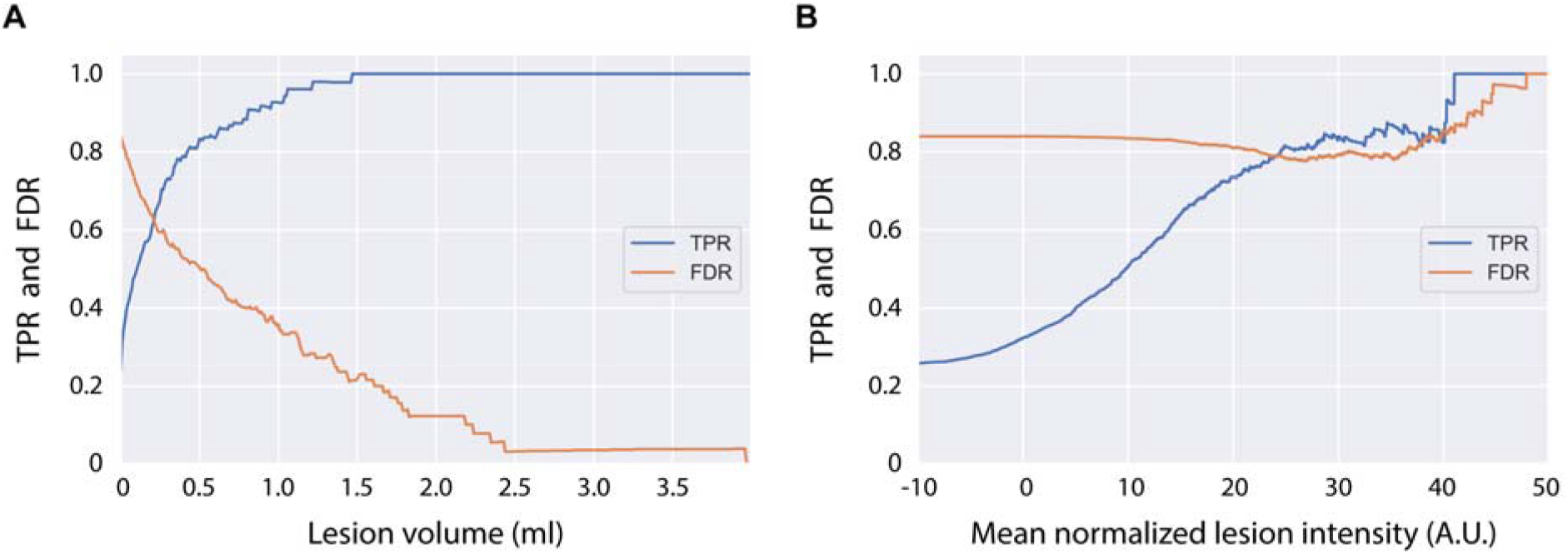
Lesion size and intensity influence detection rate. (A) The detection rate, or true positive rate (TPR), steadily increases with lesion size, with 93% detected at >1 ml, and all lesions greater than 1.5 ml being detected. The false discovery rate (FDR) decreases with lesion size, with 36% false discovery rate at >1 ml, 22% at >1.5 ml, and 3% at >2.5 ml. Though the x-axis was limited to 4 ml for illustrative purposes, lesions >20 ml were found in the dataset. (B) To analyze the relationship between lesion intensity and detection rate, image intensity values were first normalized using White Stripe (Shinohara et al. 2014). While detection rate increases as mean lesion intensity increases, the FDR remains high (>75%) across lesion intensities. The high number of false positive detections was driven by hyperintense veins and peripheral signal artifacts, as seen in figure E3.

### 3.7 Super Resolution Imaging

In one patient, a 3×4×5 mm (0.06 ml) subcortical lesion was evident near the left middle frontal gyrus on 3T (Fig. 7A) but not in any single low-field acquisition (Fig. 7D). After multi-acquisition volume averaging of 3 to 8 acquisitions, the lesion became detectable on the low-field system, and lesion intensity relative to ipsilateral white matter steadily increased with additional acquisitions. With 8 volume averages, there was a 53% increase in lesion conspicuity, which was equivalent to 72% of conspicuity at 3T (Fig. 7B). With multi-acquisition image averaging, lesions as small as 0.06 ml were discernible on 64mT scans.

**Figure 7.**
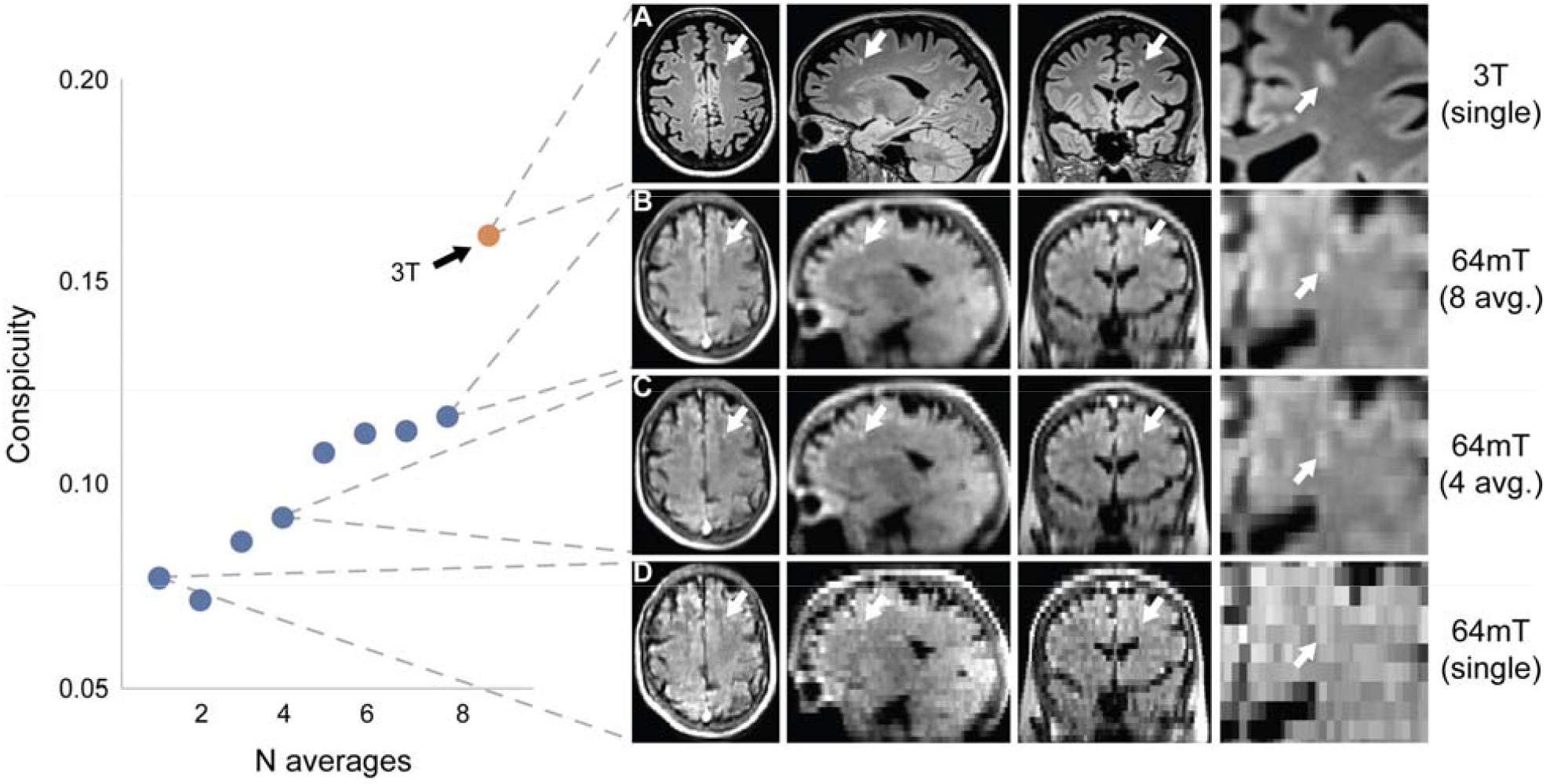
Multi acquisition image averaging increases resolution. This figure depicts a 3×4×5mm (0.06 ml) subcortical left frontal white matter lesion in a 51-55 year-old female with stable RRMS and compares 64mT FLAIR images generated from multi-acquisition image averaging to 3T imaging. The lesion is readily apparent on 3T imaging (A); however, it could not be discerned in a single 64mT acquisition (D). Volume averaging of multiple acquisitions with repositioning between scans did reveal the lesion on the low-field system (B & C). The lesion was discernible for N≥3 multi acquisition averages. The lesion was manually segmented on 3T, and the ratio of mean lesion intensity to ipsilateral adjacent white matter (WM) is given as an estimate of lesion conspicuity (red dot). In 64mT imaging, the ratio steadily increases with additional acquisition averages (blue dots). With 8 volume averages, there was a 53% increase in lesion conspicuity.

## 4. Discussion

In this study, we compared manual and automated lesion detection in paired 3T and portable 64mT brain MRI from patients with known or suspected MS at two sites. On visual inspection of 64mT images, neuroradiologists were able to detect white matter lesions in 94% (31/33) of patients with discernable 3T lesions. An automated lesion segmentation algorithm detected overlapping lesions in 91% (30/33) of patients, and estimates of total lesion burden were highly correlated between 3T and 64mT scans (r = 0.89, p < 0.001). Together, these results suggest that portable 64mT imaging could have diagnostic utility in the context of MS.

Our investigation is motivated by recent advances in hardware development and reconstruction software that address the reduced signal-to-noise and resolution associated with low-field MRI. Commercial MRI systems have predominantly trended towards higher field strengths, with low-field systems relegated to niche applications (7,11,28). Currently, there is renewed commercial interest in developing low-field MRI systems (29), including Hyperfine’s portable 64mT Swoop system, Synaptive Medical’s 0.5T Evry system, and Siemens’s 0.55T Magnetom Free.Max system, all of which have received FDA clearance since 2020. As evidence indicates that low-field systems can detect relatively subtle pathologies, such as CNS demyelination (30–32), we were motivated to investigate the sensitivity of the newly available portable 64mT MRI for MS lesions.

Our study found that lesions could be identified on 64mT scans in 94% of patients with discernible lesions at 3T. Additionally, we found a strong correlation between total lesion burden estimates on 3T and 64mT scanners. However, the smallest detected lesion size was significantly larger at 64mT (5.7 ± 1.3 mm) compared to 3T (2.1 ± 0.6 mm), indicating that smaller lesions are missed at low field. Taken together, these findings suggest that the 64mT device may be useful for tracking disease severity over time, although the device may be less suitable for making an initial diagnosis when high-field scanners are available. However, while high-field MRI will likely remain the diagnostic tool of choice in population centers of developed countries, the lower costs and infrastructure requirements of portable low-field MRI could expand clinical options for patients in low and middle income countries and rural areas (9). Within the United States, 60% of rural hospitals lack an on-site MRI (33). Additionally, most MS patients will experience mobility impairment, which can impact quality of care (34). Mobile MRI units could bring imaging to patients, providing otherwise unavailable service to sparsely populated areas and individuals who cannot travel (35,36).

The lower cost and ease of use associated with low-field MRI could also impact how early and frequently patients are imaged (37). Rovira et al. were able to predict which individuals would develop clinically definite MS using an MRI collected less than 3 months after the patient’s initial symptom onset (38). Increased scan frequency could also potentially permit earlier assessment of therapy response or detection of treatment complications, such as progressive multifocal leukoencephalopathy (16). Further studies should assess low-field MRI sensitivity for new or growing lesions over time in each clinical scenario. Our work indicates an ability to detect individual lesions at least as small as 0.06 ml using super-resolution imaging.

Low-field MRI also offers the potential to conduct large-scale studies or screening of high-risk individuals at lower cost. In MS, high-risk asymptomatic family members have an increased incidence of neurological dysfunction and neuroimaging findings associated with MS (39). Additionally, patients with radiologically isolated syndrome (i.e., patients who meet MS criteria radiologically but are clinically asymptomatic) are known to be at high risk for development of clinical MS (40). However, studies of asymptomatic individuals require large sample sizes, which cause recruitment and cost restraints. The reduced cost of low-field MRI could significantly impact the type of population based and longitudinal studies available to researchers (37).

While machine learning methods for MS lesion segmentation have yet to consistently outperform manual segmentation, they reduce the cost, time, and subjectivity associated with manual labeling (23,24). Combining low-field MRI with automated techniques can further address barriers to MRI access and image interpretation. In our work, the average Dice overlap between automated 3T and 64mT segmentations was only 0.23, with three subjects having no overlap. The low overlap was driven in part by hyperintense venous structures and peripheral artifacts on 64mT FLAIR imaging, which also resulted in a higher number of false positives (22% for lesions >1.5ml) despite comparatively high lesion sensitivity (100% for lesion >1.5 ml). Our work illustrates the importance of reassessing algorithm performance for low-field MRI sequences and indicates that retraining or tuning may be necessary to address differences in image quality and tissue contrast. In practice, an automated segmentation could serve as a biomarker for determining eligibility or endpoints in clinical trials or as a starting point for further manual refinements.

Whether gadolinium can be used to assess contrast-enhancing lesions on the 64mT device remains unknown. In our study, we saw no contrast-enhancing lesions on 3T or 64mT. Patients at site B received a standard dose of 0.1 mmol/L of a contrast agent (gadobutrol) optimized for high-field MRI; however, since 3T imaging was performed first, there was an average delay of 58 ± 21 minutes between contrast administration and 64mT imaging. At lower field strengths, the benefit from T1 shortening contrast agents is significantly reduced, and a higher gadolinium dosage (41), or potentially an alternative high-relaxivity agent (41,42), could be useful for low-field MRI. Additionally, the 58-minute delay approaches gadolinium’s half-life and this washout period can further attenuate the signal. Studies with minimal post-contrast delay and potentially higher doses of gadolinium could be conducted to better assess the potential for lesion enhancement at low field. However, even at higher doses, low-field devices may have reduced sensitivity to contrast-enhancing lesions (43).

The current study has several limitations. Our findings suggest that portable 64 mT FLAIR scans will be sensitive for white matter lesions in MS and more generally, but we focused on patients with established MS and did not assess the specificity of MS lesion detection relative to other disease processes or normal controls. In addition, sensitivity at the patient or lesion level will depend on the lesion burden and size distribution (44). We used automated 3T segmentation as ground truth, though complete labeling accuracy is challenging even at high field, and we considered lesion overlap and volume rather than lesion counts. We only evaluated a single lesion segmentation method; results may not generalize to other algorithms. Indeed, our findings indicate that both image acquisition strategies and segmentation methods can be further optimized to increase the sensitivity and accuracy of low field lesion detection for larger prospective studies. We did not assess longitudinal imaging or the ability to detect new or active lesions. Gadolinium was only administered at one of the two sites and was not administered directly for 64mT imaging. As discussed above, given that none of the patients in our cohort had gadolinium enhancing lesions on their high-field scans and the post-contrast delay before each 64mT scan, we cannot assess whether contrast enhancing lesions can be seen at 64mT.

## 5. Conclusion

In conclusion, increased imaging capabilities of low-field MRI systems warrants their re-evaluation across a range of pathologies and indications. We found that a portable 64mT scanner was sensitive to WML and that an automated algorithm designed for 3T image segmentation can be applied to 64mT data. Although additional work will be needed to evaluate portable low-field MRI systems and their capacity to carry out specific clinical functions, our findings suggest promising avenues to more accessible imaging technologies for MS around the world.

## Data Availability

The data generated in this study can be made available, with protected health information removed, upon reasonable request to the corresponding author and with a data sharing agreement between institutions in place.

https://github.com/penn-cnt/Arnold_LF-MRI_MS

## Acknowledgments

We thank the team at Hyperfine, Inc. (Guilford, CT), particularly Jonathan Rothberg, PhD, and Edward B. Welch, PhD, for technical assistance and the use of Hyperfine low-field MRI scanners. We thank the Penn Neuroradiology Research Core for assistance with patient recruitment and scanning. We also acknowledge the staff of the NINDS Neuroimmunology Clinic; Yeajin Song, MPS; Rose Cuento, CRNP; and the staff of the NIH NMR Center.

## Supplemental Material

### Study size calculation

Based on preliminary results, the maximum diameter (Dmax) of the smallest lesion on 3T imaging was estimated to be 2 mm, while Dmax was estimated to be 6 mm on 64mT imaging. The standard deviation for manual measurements was estimated to be 2 mm. Using a significance level of 0.01 and a power level of 0.95, we estimated the necessary sample size to be 18 patients to detect a significant difference between modalities (Eng 2003). Our study is well powered with over 30 patients.

---

**Figure S1.**
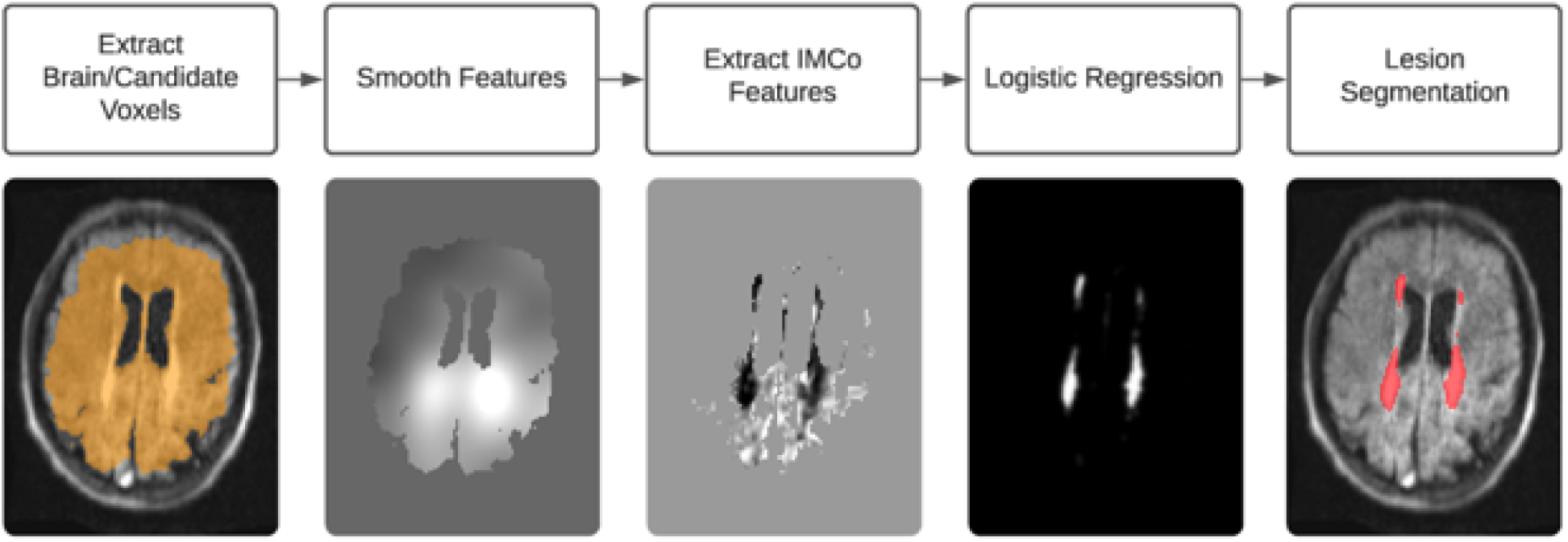
Automated segmentation algorithm. Visual description of the steps in the MIMoSA algorithm.

**Figure S2.**
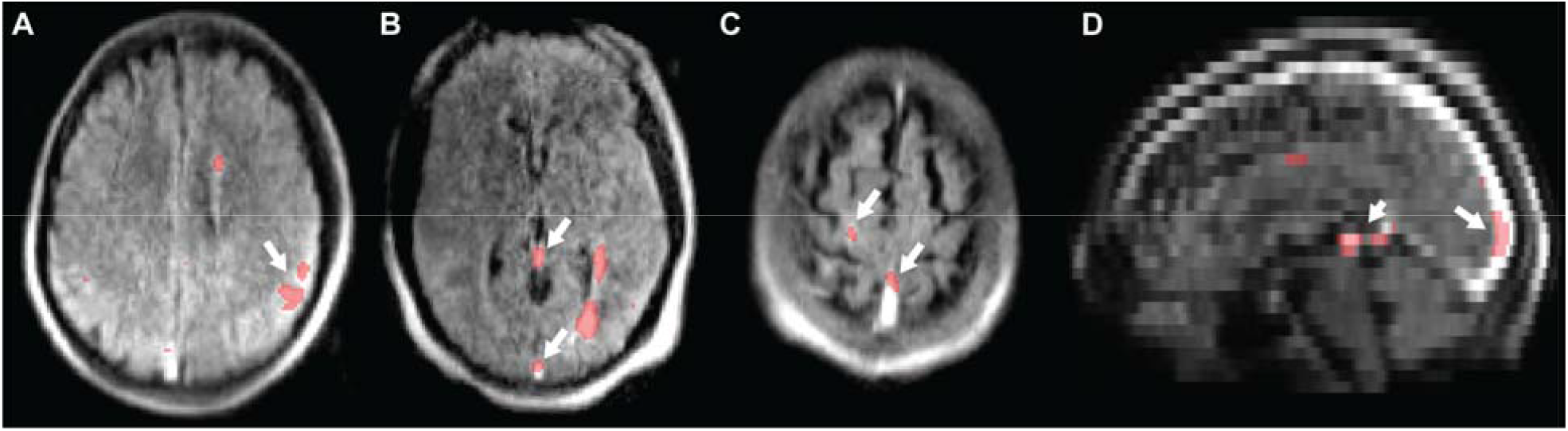
Examples of false positive lesion segmentations on 64mT T2-FLAIR. (A) False positive detections (long arrow) caused by artifactual peripheral hyperintensity in a 31-35 year-old female with RRMS. False positive detections (long arrows) caused by hyperintense venous structures as seen in (B) a 31-35 year-old female with RRMS, (C) a 51-55 year-old male with RRMS, and (D) a 46-50 year-old male with clinically isolated syndrome. A hyperintense pineal cyst (short arrow) was also a source of false positive labeling in D.

**Figure S3.**
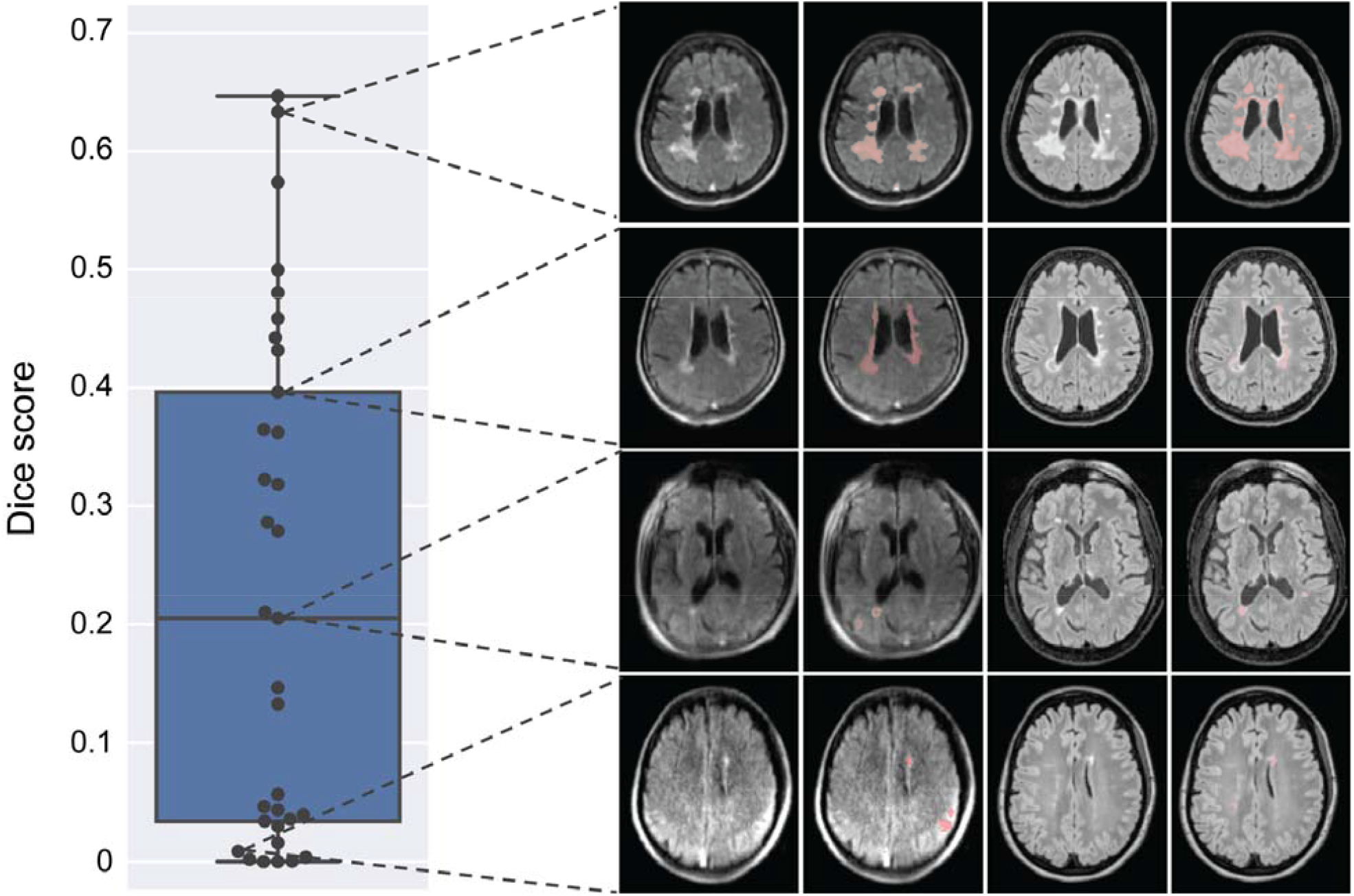
Dice score distribution of lesion segmentations. This figure highlights patients from each quartile of the Dice score distribution (mean = 0.23, standard deviation = 0.21). From left to right, the images are the original 64mT image, 64mT image with segmentation overlay, the original 3T image, and 3T image with segmentation overlay. Even patients in the lowest quartile demonstrate overlapping periventricular lesions.

**Figure S4.**
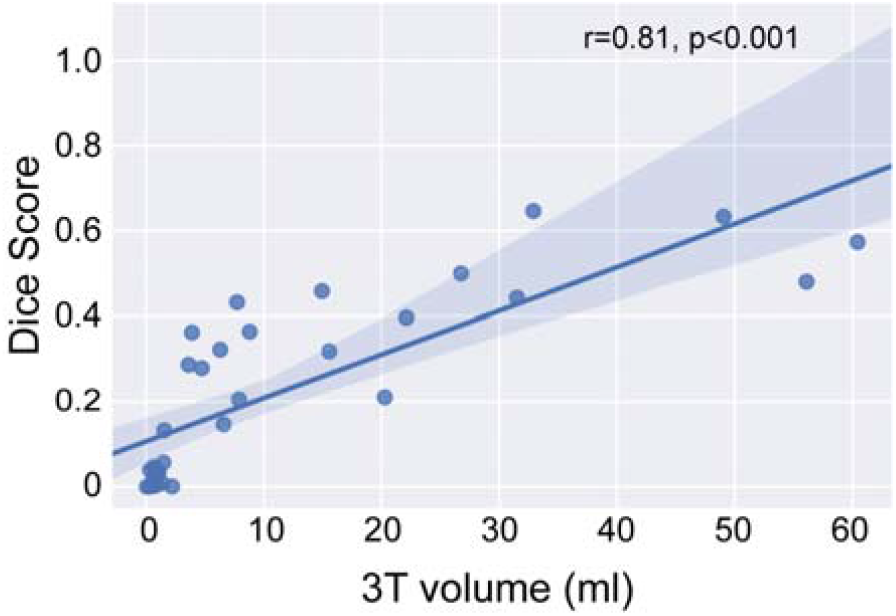
Dice score increases with lesion volume. Previous studies have found that larger lesions size are associated with higher Dice scores (25). In our study, we found a similar effect (r = 0.81, p < 0.001) such that subjects with higher lesion burden had correspondingly higher Dice scores.

## False positive detections

Visual inspection of automated 64mT segmentations revealed some consistent sources of false positive lesion detection on FLAIR images, including flow-related hyperintensities in venous structures (dural venous sinuses and small cerebral or cortical veins), other hyperintense structures such as pineal cysts, and peripheral hyperintensities due to image artifacts. Examples of these artifacts can be seen in figure E3.

In a sub-analysis, automated segmentations were manually edited in ITK-SNAP to remove some of the common false positives. In particular, we removed false positives in major sinuses (i.e., sagittal, straight, and transverse) as well as hyperintense pineal cysts. Peripheral hyperintensities caused by image artifacts or smaller cortical veins were not edited, as classification of these detections is more subjective. After manually editing the segmentations, the true positive rate (TPR) and false discovery rate (FDR) were recalculated.

There was a sharp decrease in the FDR as a function of lesion size after manual editing, reaching 11% at >1.0ml compared to 36% in the original automated segmentation (Fig. E6A). There was a modest decrease in FDR as a function of lesion intensity; however, FDR remains >65% across sensitivities (>75% in automated segmentations) (Fig. E6B). TPR remain relatively unchanged in both plots. These results highlight how optimized FLAIR sequences without hyperintense veins could decreased the rate of false positive detections by automated algorithms.

**Figure S5.**
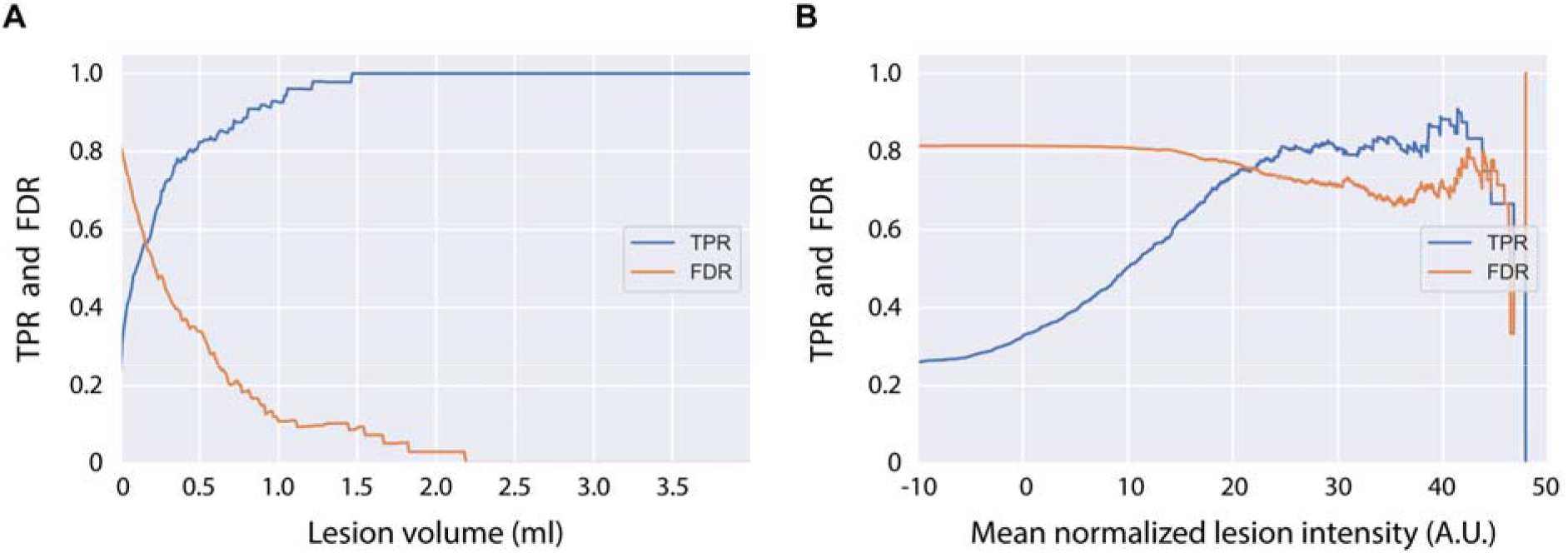
False discovery rate decreases after removing hyperintense veins. After manually removing hyperintense veins and other hyperintense brain structures from segmentations, the false discovery rate (FDR) decreases. (A) The FDR decreases as a function of lesion size. Manually edited segmentations had a lower FDR relative to the unedited automated segmentations (>1 ml: unedited=36%, edited=11%, >1.5ml: unedited=22%, edited=9%, >2.5ml: unedited=3%, edited=0%). (B) When analyzing FDR as a function of mean lesion intensity, the effect was less pronounced. In unedited segmentations, FDR was high (>75%) across lesion intensities. While there was a modest reduction after manual editing, FDR remained >65%.

## References

1. Reich DS, Lucchinetti CF, Calabresi PA. Multiple Sclerosis. Longo DL, editor. N Engl J Med. N Engl J Med; 2018;378(2):169–180. doi: 10.1056/NEJMra1401483.

2. Thompson AJ, Banwell BL, Barkhof F, et al. Diagnosis of multiple sclerosis: 2017 revisions of the McDonald criteria. Lancet Neurol. Lancet Publishing Group; 2018. p. 162–173. doi: 10.1016/S1474-4422(17)30470-2.

3. Filippi M, Preziosa P, Banwell BL, et al. Assessment of lesions on magnetic resonance imaging in multiple sclerosis: practical guidelines. Brain. Oxford University Press; 2019. p. 1858–1875. doi: 10.1093/brain/awz144.

4. Noyes K, Weinstock-Guttman B. Impact of diagnosis and early treatment on the course of multiple sclerosis. Am. J. Manag. Care. Ascend Media; 2013. p. s321–31. https://europepmc.org/article/med/24494633. accessed October 18, 2021.

5. Wallin MT, Culpepper WJ, Campbell JD, et al. The prevalence of MS in the United States: A population-based estimate using health claims data. Neurology. Lippincott Williams and Wilkins; 2019;92(10):E1029–E1040. doi: 10.1212/WNL.0000000000007035.

6. Tullman MJ. Overview of the epidemiology, diagnosis, and disease Progression Associated With multiple Sclerosis. Am J Manag Care. 2013;19:S15–S20.

7. Marques JP, Simonis FFJ, Webb AG. LowlJfield MRI: An MR physics perspective. J Magn Reson Imaging. John Wiley and Sons Inc.; 2019;49(6):1528–1542. doi: 10.1002/jmri.26637.

8. Ogbole GI, Adeyomoye AO, Badu-Peprah A, Mensah Y, Nzeh DA. Survey of magnetic resonance imaging availability in West Africa. Pan Afr Med J. African Field Epidemiology Network; 2018;30. doi: 10.11604/pamj.2018.30.240.14000.

9. Maru DS-R, Schwarz R, Andrews J, Basu S, Sharma A, Moore C. Turning a blind eye: the mobilization of radiology services in resource-poor regions. Global Health. 2010;6(1):18. doi: 10.1186/1744-8603-6-18.

10. Wald LL, McDaniel PC, Witzel T, Stockmann JP, Cooley CZ. Low-cost and portable MRI. J Magn Reson Imaging. John Wiley and Sons Inc.; 2020;52(3):686–696. doi: 10.1002/jmri.26942.

11. Campbell-Washburn AE, Ramasawmy R, Restivo MC, et al. Opportunities in Interventional and Diagnostic Imaging by Using High-Performance Low-Field-Strength MRI. Radiology. Radiological Society of North America Inc.; 2019;293(2):384–393. doi: 10.1148/radiol.2019190452.

12. Danni TU, Goyal MS, Dworkin JD, et al. Multi-modal biomarkers of cerebral edema in low resolution MRI. bioRxiv. Cold Spring Harbor Laboratory; 2020. p. 2020.12.23.424020. doi: 10.1101/2020.12.23.424020.

13. Sheth KN, Mazurek MH, Yuen MM, et al. Assessment of brain injury using portable, low-field magnetic resonance imaging at the bedside of critically ill patients. JAMA Neurol. American Medical Association; 2020;E1–E7. doi: 10.1001/jamaneurol.2020.3263.

14. Mazurek MH, Cahn BA, Yuen MM, et al. Portable, bedside, low-field magnetic resonance imaging for evaluation of intracerebral hemorrhage. Nat Commun. Nature Publishing Group; 2021;12(1):5119. doi: 10.1038/s41467-021-25441-6.

15. Turpin J, Unadkat P, Thomas J, et al. Portable Magnetic Resonance Imaging for ICU Patients. Crit Care Explor. Ovid Technologies (Wolters Kluwer Health); 2020;2(12):e0306. doi: 10.1097/cce.0000000000000306.

16. Scarpazza C, Signori A, Cosottini M, Sormani MP, Gerevini S, Capra R. Should frequent MRI monitoring be performed in natalizumab-treated MS patients? A contribution to a recent debate. Mult Scler J. SAGE Publications Ltd; 2020;26(10):1227–1236. doi: 10.1177/1352458519854162.

17. Yushkevich PA, Piven J, Hazlett HC, et al. User-guided 3D active contour segmentation of anatomical structures: Significantly improved efficiency and reliability. Neuroimage. Academic Press; 2006;31(3):1116–1128. doi: 10.1016/J.NEUROIMAGE.2006.01.015.

18. Tustison NJ, Avants BB, Cook PA, et al. N4ITK: Improved N3 Bias Correction. IEEE Trans Med Imaging. 2010;29(6):1310–1320. doi: 10.1109/TMI.2010.2046908.

19. Tustison NJ, Cook PA, Holbrook AJ, et al. The ANTsX ecosystem for quantitative biological and medical imaging. Sci Rep. NLM (Medline); 2021;11(1):9068. doi: 10.1038/s41598-021-87564-6.

20. Avants, Brian B., Tustison, Nick, Song G. Advanced normalization tools (ANTS). Insight j 2. 2009;1–35.

21. Doshi J, Erus G, Ou Y, Gaonkar B, Davatzikos C. Multi-Atlas Skull-Stripping. Acad Radiol. Elsevier; 2013;20(12):1566–1576. doi: 10.1016/j.acra.2013.09.010.

22. Shinohara RT, Sweeney EM, Goldsmith J, et al. Statistical normalization techniques for magnetic resonance imaging. NeuroImage Clin. Elsevier Inc.; 2014;6:9–19. doi: 10.1016/j.nicl.2014.08.008.

23. Valcarcel AM, Linn KA, Vandekar SN, et al. MIMoSA: An Automated Method for Intermodal Segmentation Analysis of Multiple Sclerosis Brain Lesions. J Neuroimaging. Blackwell Publishing Inc.; 2018;28(4):389–398. doi: 10.1111/jon.12506.

24. Valcarcel AM, Linn KA, Khalid F, et al. A dual modeling approach to automatic segmentation of cerebral T2 hyperintensities and T1 black holes in multiple sclerosis. NeuroImage Clin. Elsevier Inc.; 2018;20:1211–1221. doi: 10.1016/j.nicl.2018.10.013.

25. Oguz I, Carass A, Pham DL, et al. Dice overlap measures for objects of unknown number: Application to lesion segmentation. Lect Notes Comput Sci (including Subser Lect Notes Artif Intell Lect Notes Bioinformatics). Springer Verlag; 2018. p. 3–14. doi: 10.1007/978-3-319-75238-9_1.

26. Boudraa AO, Dehak SMR, Zhu YM, Pachai C, Bao YG, Grimaud J. Automated segmentation of multiple sclerosis lesions in multispectral MR imaging using fuzzy clustering. Comput Biol Med. Elsevier Science Ltd; 2000;30(1):23–40. doi: 10.1016/S0010-4825(99)00019-0.

27. Jovicich J, Czanner S, Han X, et al. MRI-derived measurements of human subcortical, ventricular and intracranial brain volumes: Reliability effects of scan sessions, acquisition sequences, data analyses, scanner upgrade, scanner vendors and field strengths. Neuroimage. Academic Press; 2009;46(1):177–192. doi: 10.1016/j.neuroimage.2009.02.010.

28. O’Reilly T, Teeuwisse WM, Gans D, Koolstra K, Webb AG. In vivo 3D brain and extremity MRI at 50 mT using a permanent magnet Halbach array. Magn Reson Med. John Wiley and Sons Inc; 2021;85(1):495–505. doi: 10.1002/mrm.28396.

29. Sarracanie M, Salameh N. Low-Field MRI: How Low Can We Go? A Fresh View on an Old Debate. Front Phys. Frontiers Media S.A.; 2020;8:172. doi: 10.3389/fphy.2020.00172.

30. Mateen FJ, Cooley CZ, Stockmann JP, Rice DR, Vogel AC, Wald LL. Low-field Portable Brain MRI in CNS Demyelinating Disease. Mult Scler Relat Disord. Elsevier; 2021;102903. doi: 10.1016/j.msard.2021.102903.

31. Orrison WW, Stimac GK, Stevens EA, et al. Comparison of CT, low-field-strength MR imaging, and high-field-strength MR imaging. Work in progress. Radiology. Radiology; 1991. p. 121–127. doi: 10.1148/radiology.181.1.1887020.

32. Arnold TC, Baldassano SN, Litt B, Stein JM. Simulated diagnostic performance of low-field MRI: Harnessing open-access datasets to evaluate novel devices. Magn Reson Imaging. Elsevier; 2021; doi: 10.1016/j.mri.2021.12.007.

33. Ginde AA, Foianini A, Renner DM, Valley M, Camargo CA. Availability and quality of computed tomography and magnetic resonance imaging equipment in U.S. emergency departments. Acad Emerg Med. Acad Emerg Med; 2008;15(8):780–783. doi: 10.1111/j.1553-2712.2008.00192.x.

34. Sutliff MH. Contribution of impaired mobility to patient burden in multiple sclerosis. Curr. Med. Res. Opin. Curr Med Res Opin; 2010. p. 109–119. doi: 10.1185/03007990903433528.

35. Emerging Ethical Issues Raised by Highly Portable MRI Research in Remote and Resource-Limited International SettingsShen FX, Wolf SM, Bhavnani S, et al. Emerging Ethical Issues Raised by Highly Portable MRI Research in Remote and Resource-Limited International Settings. Neuroimage. Academic Press; 2021;118210. doi: 10.1016/j.neuroimage.2021.118210.

36. Deoni SC, Deoni AT, Burton P, et al. Residential MRI: Development of A Mobile Anywhere-Everywhere MRI Lab. 2021; doi: 10.21203/rs.3.rs-1121934/v1.

37. Deoni SCL, D’Sa V, Volpe A, et al. Remote and At-Home Data Collection: Considerations for the NIH HEALthy Brain and Cognitive Development (HBCD) Study. Dev Cogn Neurosci. Elsevier; 2022;101059. doi: 10.1016/j.dcn.2022.101059.

38. Rovira À, Swanton J, Tintoré M, et al. A single, early magnetic resonance imaging study in the diagnosis of multiple sclerosis. Arch Neurol. Arch Neurol; 2009;66(5):587–592. doi: 10.1001/archneurol.2009.49.

39. Xia Z, Steele SU, Bakshi A, et al. Assessment of early evidence of multiple sclerosis in a prospective study of asymptomatic high-risk family members. JAMA Neurol. American Medical Association; 2017;74(3):293–300. doi: 10.1001/jamaneurol.2016.5056.

40. Hosseiny M, Newsome SD, Yousem DM. Radiologically isolated syndrome: A review for neuroradiologists. Am. J. Neuroradiol. American Society of Neuroradiology; 2020. p. 1542–1549. doi: 10.3174/ajnr.A6649.

41. Desai NK, Runge VM. Contrast Use at Low Field: A Review. Top Magn Reson Imaging. 2003;14(5):360–364. doi: 10.1097/00002142-200310000-00002.

42. Bendszus M, Roberts D, Kolumban B, et al. Dose Finding Study of Gadopiclenol, a New Macrocyclic Contrast Agent, in MRI of Central Nervous System. Invest Radiol. Lippincott Williams and Wilkins; 2020;55(3):129–137. doi: 10.1097/RLI.0000000000000624.

43. Ertl-Wagner B, Reith W, Sartor K. Low field-low cost: Can low-field magnetic resonance systems replace high-field magnetic resonance systems in the diagnostic assessment of multiple sclerosis patients? Eur Radiol. Springer; 2001;11(8):1490–1494. doi: 10.1007/s003300000806.

44. Commowick O, Istace A, Kain M, et al. Objective Evaluation of Multiple Sclerosis Lesion Segmentation using a Data Management and Processing Infrastructure. Sci Rep. Nature Publishing Group; 2018;8(1). doi: 10.1038/s41598-018-31911-7.

## References

1. Eng J. Sample size estimation: how many individuals should be studied? Radiology. 2003 May;227(2):309-13. Doi: 10.1148/radiol.2272012051. PMID: 12732691.

